# Research on prevention and control strategies for norovirus clusters: An analysis of epidemiological characteristics and influencing factors of norovirus clusters in Hangzhou, China

**DOI:** 10.1101/2025.09.23.25336522

**Authors:** Song Kai, Wang Jing, Duan Xiaojian, Huo Liangliang, Ren Xiaobin, Wang Jiayun

**Affiliations:** Hangzhou Center for Disease Control and Prevention ( Hangzhou Health Supervision Institution), Hangzhou, China. 310021; Institute of Infectious Disease Control and Prevention

**Keywords:** norovirus, epidemiological characteristics, school, prevention and control

## Abstract

Norovirus is currently one of the leading pathogens of infectious diarrhea, imposing a substantial disease burden annually. In recent years, Hangzhou has experienced a year-by-year increase in norovirus clusters across diverse settings, including schools, hospitals, nursing homes, corporations, restaurants, hotels, villages, and construction sites. Individual clusters often affect dozens to hundreds of individuals, indicating persistent gaps in certain aspects of prevention and control efforts.

Under constraints of funding and human resources, public health authorities must identify factors warranting prioritized intervention based on their socioeconomic impact. We analyzed 432 norovirus clusters in Hangzhou from 2018 to 2023 and identified transmission route, urban-rural distribution, setting type, and genogroup as key determinants of outbreak scale.The high-incidence period occurs from October to May, during which prevention and control measures should be enhanced. Primary schools and kindergartens are the main settings, with 6.85% of primary schools and 2.63% of kindergartens reporting such clusters. These should be prioritized for targeted interventions. Person-to-person transmission is the dominant route. Strengthening routine environmental disinfection, improving ventilation, and promoting correct handwashing practices among students are essential. Although less common, foodborne transmission leads to larger outbreaks, with more cases, longer duration, and broader classroom impact. Thus, enhanced hygiene supervision of school canteens is critical.Suburban and rural areas, often facing economic and cultural disadvantages, require increased allocation of prevention resources and tailored public health support.

## 1. Introduction

With the development of the global economic and social, viruses have replaced bacteria and parasites as the leading pathogens of infectious diarrhea, with norovirus (Norovirus, NoV) being one of the primary causative agents. A meta-analysis spanning 1997–2021 and incorporating data from 72 countries revealed that approximately 16% of acute gastroenteritis cases over the past two decades were attributable to norovirus infection [1]. Particularly following the widespread administration of rotavirus vaccines, the epidemiological significance of norovirus has become even more pronounced, imposing a substantial disease burden [2]. In recent years, clusters of norovirus in China have exhibited an increasing annual trend, with affected regions expanding [3]. Similarly, Hangzhou has experienced a notable rise in norovirus clusters, occurring in diverse settings such as schools, hospitals, nursing homes, corporate offices, restaurants, hotels, villages, and construction sites. These clusters frequently result in dozens to hundreds of cases, with schools being the most common setting, drawing growing attention from parents and media. Consequently, relevant authorities have repeatedly issued updated requirements for norovirus prevention and control in schools.

The severity of the outbreak situation highlights persistent gaps in certain aspects of prevention and control efforts. As there are currently no approved vaccines against norovirus nor specific treatments for infection, prevention and outbreak containment remain critically important. However, under constraints of funding and human resources, disease prevention and control agencies must identify the most socioeconomically impactful factors for prioritized intervention from vast amounts of surveillance data. This study analyzes data from norovirus cluster outbreaks in Hangzhou from 2018 to 2023 to inform more targeted and effective prevention and control strategies.

## 2. Methods

### 2.1 Data source

The data were obtained from the China Information System for Disease Control and Prevention and field investigations of outbreak responses. The study included norovirus acute gastroenteritis clusters with onset dates of the first cases between January 1, 2018, and December 31, 2023.All cases underwent detailed epidemiological investigation, including demographic characteristics, onset time, clinical symptoms, disappearance time of symptoms, medical treatment situation, sampling time, test results, etc.). Concurrently, investigations covered baseline information of outbreak settings (e.g., schools, and workplaces) and potential risk factors such as food, drinking water, and environmental surfaces. Anal swab samples were collected from some cases and high-risk groups (such as teachers and cafeteria workers). Anal swab specimens were detected for the norovirus by municipal or county-level CDC laboratories using PCR-based methods. The number of schools in the study area was extracted from the Hangzhou Statistical Yearbook published by the Hangzhou Statistics Bureau.

### 2.2 Definitions and Classifications

#### 2.2.1 Case

During norovirus-associated outbreaks or clusters, individuals meeting the case definition (≥3 loose/watery stools and/or ≥2 vomiting episodes within 24 hours) with epidemiological links to laboratory-confirmed cases were identified.

#### 2.2.2 Cluster/Outbreak

According to the Technical Guidelines for Norovirus Outbreak Investigation and Prevention in China (2015 Edition) [4]:

**Cluster**: ≥5 epidemiologically linked norovirus infections (including ≥2 laboratory-confirmed cases) occurring within 3 days in the same institutional setting (e.g., school, childcare facility, hospital, nursing home, factory, construction site, cruise ship, or community/village).

**Outbreak**: ≥20 epidemiologically linked norovirus infections (including ≥2 laboratory-confirmed cases) occurring within 7 days in the same institutional setting (as above).

For clarity, cluster cases not meeting the outbreak threshold were termed **“non-outbreak clusters”** in this study.

#### 2.2.2 Urban-Rural Classification

Districts were categorized as:

Urban: Shangcheng, Gongshu, Xihu, Binjiang, Qiantang, and mingsheng.

Rural: Xiaoshan, Yuhang, Linping, Fuyang, Jiande, Tonglu, Chun’an, and Lin’an.

#### 2.2.3 Duration

Calculated as the interval (in decimal days) between Symptom onset in the index case and Symptom resolution in the terminal case.

#### 2.2.4 Reporting Timeliness

Defined as the time difference (in days) between official outbreak reporting time and symptom onset in the index case.

#### 2.2.5 School Classification

In China, children typically begin kindergarten at age 3. Primary school usually lasts six years, followed by three years each of junior high and senior high school. Therefore, the age ranges are as follows: Kindergarten(3–5 years), Primary school(6–12 years), Junior high school(13–15 years), Senior high school(16–18 years).

#### 2.2.6 Attack Rate

**Attack rate = (Number of cases / Number of persons at risk) × 100%**

For person-to-person transmission outbreaks, which typically affected limited classrooms, the denominator included the total number of students in involved classes. For foodborne outbreaks, which often spread throughout the school, the denominator was the total school population.

### 2.3 Statistical Analysis

Data were collated and analyzed using Excel 2010 and SPSS 19.0. Descriptive epidemiology methods characterized the temporal, spatial, and population distributions of clusters. Continuous variables were reported as median (M) with interquartile range (Q1, Q3). Group comparisons employed Mann-Whitney U test (two-group comparisons), Kruskal-Wallis H test (multi-group comparisons) and Chi-square test (categorical variables). Multivariable analysis used unconditional logistic regression with significance set at α=0.05 (two-tailed).

## 3. Results

### 3.1 General Characteristics

From 2018 to 2023, Hangzhou reported a total of 432 norovirus acute gastroenteritis clusters with a median of 12 cases per cluster (IQR 9-18) and a median attack rate of 22.50% (IQR 12.87%-34.38%), accumulating 6,904 cases including 2,613 laboratory-confirmed cases (37.85%) with no fatal cases. Among these, 89 clusters (20.60%) met the outbreak criteria, showing a median of 27 cases per outbreak (IQR 22-39) with a median attack rate of 25.00% (IQR 16.67%-35.48%), totaling 3,079 cases including 745 laboratory-confirmed cases (20.09%). Genogroup analysis revealed GⅡ as the predominant strain (355 clusters, 82.18%), followed by GI (59 clusters, 13.66%), while GI and GⅡ co-infections were least frequent (18 clusters, 4.17%), as detailed in Table 1.

**Table 1.** Basic characteristics of norovirus clusters in Hangzhou, 2018-2023.

| year | Total | Non-outbreak clusters n(%) | Outbreak clusters n(%) | Urban/Rural distribution(%) |  | Transmission route n(%) |  |  | Setting n(%) |  |  |  |  | Genogroup distribution n(%) |  |  |
| --- | --- | --- | --- | --- | --- | --- | --- | --- | --- | --- | --- | --- | --- | --- | --- | --- |
|  |  |  |  | Urban | Rural | Person-to-person | Foodborne | Waterborne | Kindergarten | Primary school | Junior high | Senior high | Other | GI | GII | Mixed |
| 2018 | 38 | 33(86.84) | 5(13.16) | 27(71.05) | 11(28.95) | 37(97.37) | 1(2.63) | 0(0.00) | 13(34.21) | 20(52.63) | 2(5.26) | 2(5.26) | 1(2.63) | 1(2.63) | 34(89.47) | 3(7.90) |
| 2019 | 94 | 74(78.72) | 20(21.28) | 60(63.83) | 34(36.17) | 82(87.23) | 11(11.7) | 0(0.00) | 20(21.28) | 57(60.64) | 6(6.38) | 7(7.45) | 4(4.26) | 28(29.79) | 61(64.89) | 5(5.32) |
| 2020 | 125 | 97(77.60) | 28(22.40) | 63(50.40) | 62(49.60) | 116(92.80) | 9(7.20) | 0(0.00) | 53(42.40) | 56(44.80) | 3(2.40) | 7(5.60) | 6(4.80) | 6(4.80) | 118(94.40) | 1(0.80) |
| 2021 | 74 | 62(83.78) | 12(16.22) | 52(70.27) | 22(29.73) | 68(91.89) | 4(5.41) | 1(1.35) | 37(50) | 27(36.49) | 1(1.35) | 2(2.70) | 7(9.46) | 7(9.46) | 65(87.84) | 2(2.70) |
| 2022 | 30 | 23(76.67) | 7(23.33) | 25(83.33) | 5(16.67) | 29(96.67) | 1(3.33) | 0(0.00) | 15(50.00) | 13(43.33) | 1(3.33) | 0(0.00) | 1(3.33) | 9(30.00) | 17(56.67) | 4(13.33) |
| 2023 | 71 | 54(76.06) | 17(23.94) | 40(56.34) | 31(43.66) | 66(92.96) | 5(7.04) | 0(0.00) | 27(38.03) | 31(43.66) | 4(5.63) | 5(7.04) | 4(5.63) | 8(11.27) | 60(84.51) | 3(4.23) |
| Total | 432 | 343(79.40) | 89(20.60) | 267(61.81) | 165(38.19) | 400(92.59) | 31(7.18) | 1(0.23) | 165(38.19) | 204(47.22) | 17(3.94) | 23(5.32) | 23(5.32) | 59(13.66) | 355(82.18) | 18(4.17) |

### 3.2 Epidemiological Characteristics

#### 3.2.1 Seasonal Distribution

Norovirus acute gastroenteritis clusters in this municipality predominantly occurred from October to May, with peak incidence observed in November and December. Case numbers declined significantly during February (typically coinciding with school winter vacations in China), followed by a minor resurgence in March after schools reopened, attributed to approximately one week of viral transmission. Cluster frequency progressively decreased with rising temperatures, reaching the annual nadir between June and August.

Notably, GI genogroup clusters exhibited distinct seasonality, lacking the winter peak but demonstrating a minor surge during April-May. Mixed GI/GII infections showed no clear temporal clustering pattern due to limited case numbers.

These findings correlated with norovirus detection patterns among “other infectious diarrhea” cases reported through the China Disease Control and Prevention Information System, where monthly distribution trends mirrored cluster activity (Figure 1).

**Fig 1.**
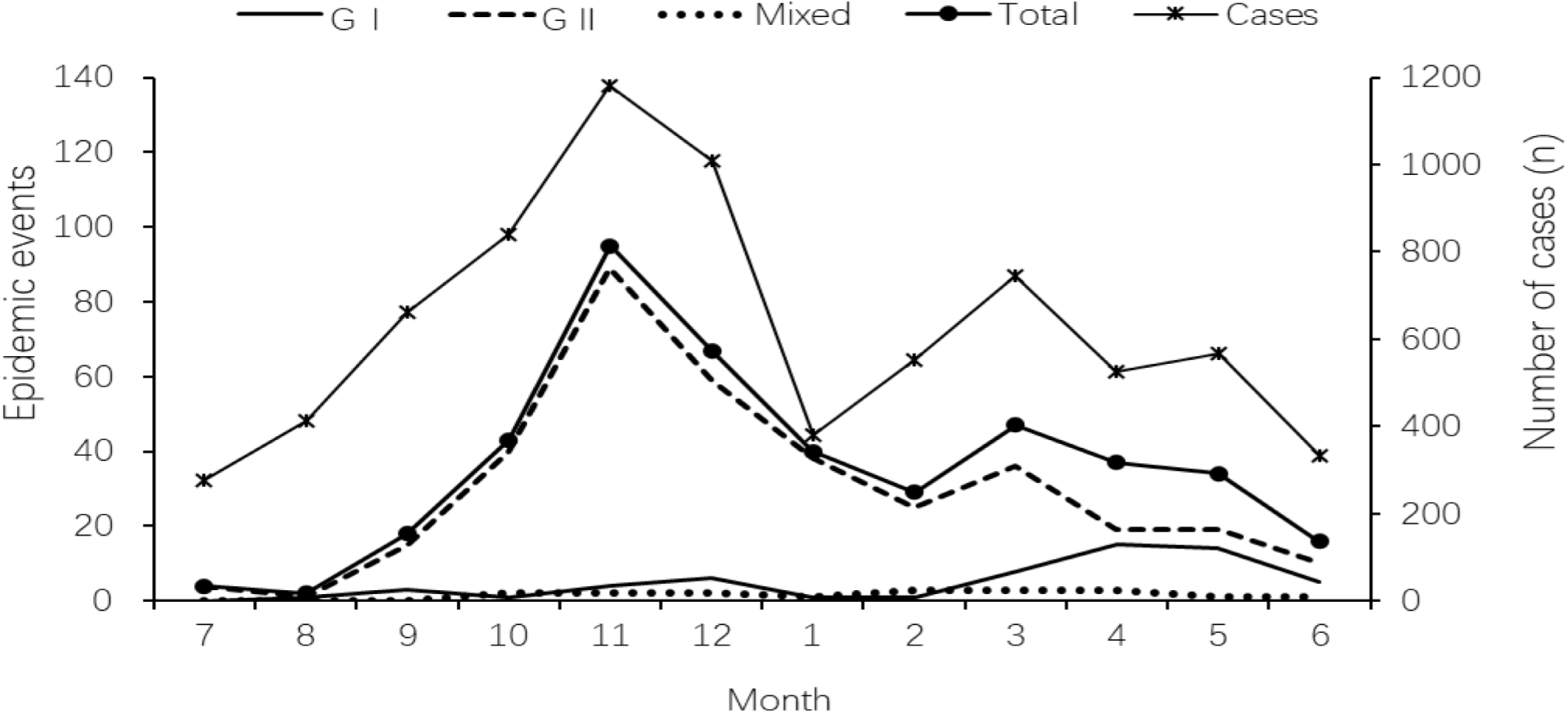
Monthly distribution of norovirus clusters by different genogroups. “Case number” refers to the count of norovirus-positive cases reported as “other infectious diarrhea diseases” in the China Disease Control and Prevention Information System. Since the high-incidence period extends continuously from October to May of the following year, the x-axis in this figure begins in July to more clearly display the continuous months of high activity. The number of clusters decreased significantly during the winter vacation period, which generally occurs in the latter half of January and the first half of February.

#### 3.2.2 Geographic and Institutional Distribution

The top five districts/counties by cumulative case reports were Binjiang District (70 clusters, 16.20%), Xiaoshan District (69 clusters, 15.97%), Qiantang District (60 clusters, 13.89%), Shangcheng District (57 clusters, 13.19%), and Gongshu District (49 clusters, 11.34%), collectively accounting for 70.60% (305/432) of total clusters. Urban areas reported 267 clusters (61.81%) compared to 165 clusters (38.19%) in suburban/rural regions.

Primary schools (47.22%, 204/432) and kindergartens (38.19%, 165/432) were the predominant outbreak settings, comprising 85.41% (369/432) of total clusters. Educational institutions (kindergartens, primary, junior high, and senior high schools) accounted for 94.68% (409/432), with 23 clusters (5.32%) occurring in other settings: universities (5), training institutions (4), hotels/restaurants (4), corporations (3), villages (2), elderly care facilities (2), hospitals (1), athletic training centers (1), and construction sites (1).

The six-year average incidence rates revealed: Highest in primary schools (6.85%, 204/2978), Intermediate in kindergartens (2.63%, 165/6280) and senior high schools (2.66%, 23/865),Lowest in junior high schools (0.99%, 17/1721).Urban-rural comparisons showed statistically significant differences (χ² tests):Higher incidence in urban kindergartens (χ²=50.075, P<0.01) and primary schools (χ²=80.447, P<0.01), Lower incidence in urban senior high schools (χ²=4.097, P<0.05), No significant difference for junior high schools (χ²=1.039, P>0.05).(See Table 2 for detailed comparisons).

**Table 2.** Urban-rural distribution of norovirus gastroenteritis clusters in educational settings, Hangzhou (2018-2023)

|  | Kindergarten |  |  |  |  | Primary School |  |  |  |  | Junior High |  |  |  |  | Senior High |  |  |  |  |
| --- | --- | --- | --- | --- | --- | --- | --- | --- | --- | --- | --- | --- | --- | --- | --- | --- | --- | --- | --- | --- |
| | n | N | % | $\chi^2$ | P | n | N | % | $\chi^2$ | P | n | N | % | $\chi^2$ | P | n | N | % | $\chi^2$ | P |
| Urban | 110 | 2514 | 4.38 | 50.075 | <0.01 | 131 | 1050 | 12.48 | 80.447 | <0.01 | 9 | 703 | 1.28 | 1.039 | 0.308 | 5 | 366 | 1.37 | 4.097 | 0.043 |
| Rural | 55 | 3766 | 1.46 |  |  | 73 | 1928 | 3.79 |  |  | 8 | 1018 | 0.79 |  |  | 18 | 499 | 3.61 |  |  |
| Total | 165 | 6280 | 2.63 |  |  | 204 | 2978 | 6.85 |  |  | 17 | 1721 | 0.99 |  |  | 23 | 865 | 2.66 |  |  |

Note: n represents the number of norovirus cluster events during 2018-2023; N represents the cumulative sum of annual school numbers for each category during 2018-2023.

### 3.3 Transmission Pathways

The norovirus acute gastroenteritis clusters in this region were predominantly transmitted through person-to-person contact (400/432, 92.59%), with fewer cases of foodborne transmission (31/432, 7.18%) and only one waterborne incident (0.23%, occurring in a village due to contamination of a stream used for daily washing). Foodborne transmission clusters had significantly higher progression to outbreak status (90.32%, 28/31) compared to person-to-person transmission clusters (15%, 60/400; χ²=100.455, P<0.01) Urban areas showed:94.76% (253/267) person-to-person transmission, 5.24% (14/267) foodborne transmission.Suburban/rural areas demonstrated:89.09% (147/165) person-to-person transmission,10.30% (17/165) foodborne transmission,0.61% (1/165) waterborne transmission.The higher proportion of foodborne transmission in suburban/rural areas was statistically significant (χ²=3.919, P<0.01).Senior high schools had markedly higher foodborne transmission rates (56.52%, 13/23) compared to: Kindergartens (0.00%, 0/165),Primary schools (5.39%, 11/204),Junior high schools (5.88%, 1/17),(All differences statistically significant).

### 3.4 Genogroup distribution

GII was predominant (82.18%, 355/432),GI accounted for 13.66% (59/432),Mixed infections represented 4.17% (18/432),Temporal variation: GI prevalence peaked in 2019 (29.79%) and 2022 (30.00%) (Figure 2).School-level differences: GI was more common in primary schools (20.59%), mixed infections showed an increasing trend with higher school grade levels.Genotyping results (12 samples):GII.7 was dominant (58.33%, 7/12),GII.3 and GII.17: 2 cases each,GII.4: 1 case,Suggests GII.7 was likely the predominant circulating genotype during the study period.These findings suggest distinct epidemiological patterns between GI and GII genogroups, as illustrated in Figure 2.

**Fig 2.**
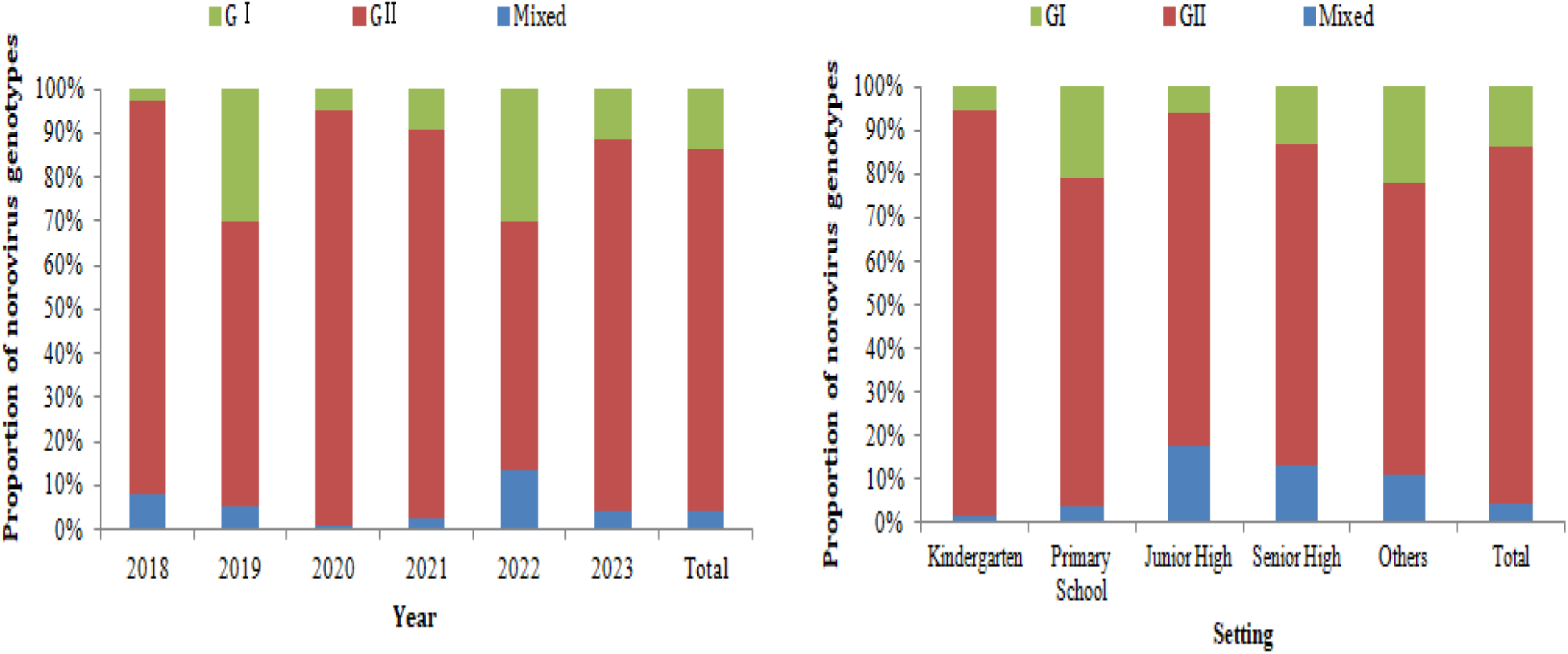
Genogroup distribution of norovirus outbreak strains in Hangzhou, 2018–2023. This figure illustrates the distribution of norovirus genogroups across different years and settings. The proportions of each genogroup fluctuated over time, with GII remaining the predominant genotype overall.

### 3.5 Outbreak Scale Characteristics

The median (IQR) values for overall norovirus clusters were: 12 (9-18) cases per event, 22.56% (13.33%-34.36%) attack rate, and 2.21 (1.17-4.07) days duration. Outbreak-status clusters showed higher medians: 27 (22-38.75) cases, 11.38% (4.00%-22.60%) attack rate, and 4.64 (2.15-8.32) days duration.

Transmission route significantly influenced outbreak characteristics. Person-to-person transmission typically affected 1-3 classes (median 12 cases, IQR 8-17; attack rate 24%, IQR 15.21%-34.38%; duration 2.02 days, IQR 1.08-3.44), with 67.45% (259/384) confined to single classrooms. In contrast, foodborne outbreaks (median 39 cases, IQR 27-63; attack rate 3.34%, IQR 1.64%-4.54%; duration 7.17 days, IQR 4.34-9.67) showed wider dispersion (median 16 classes, IQR 10-24), with 44% (11/25) affecting >20 classes (Table 3). All scale parameters were significantly greater for foodborne versus person-to-person transmission (p<0.001).

**Table 3.** Epidemiological characteristics of norovirus clusters in Hangzhou, 2018-2023.

|  | Cluster count<br>(%) | Cases per cluster<br>M(Q1,Q3) | Classes affected<br>M(Q1,Q3) | Reporting timeliness(days)<br>M(Q1,Q3) | Cluster duration(days)<br>M(Q1,Q3) | Attack rate (%)<br>M(Q1,Q3) |
| --- | --- | --- | --- | --- | --- | --- |
| **Person-to-person transmission** | 400(92.59) | 12(8,17) | 1(1,2) | 2.02(1.10,3.03) | 2.02(1.08,3.44) | 24.00(15.21,34.98) |
| Urban-rural |  |  |  |  |  |  |
| Urban | 253(63.25) | 11(8,16) | 1(1,1) | 2.08(1.17,3.05) | 2.00(1.00,3.05) | 24.64(17.11,35.81) |
| Rural | 147(36.75) | 13(10,18) | 1(1,3) | 2.00(1.00,3.00) | 2.19(1.38,5.04) | 21.67(12.50,34.78) |
| Settings |  |  |  |  |  |  |
| Kindergartens | 165(41.25) | 10(8,13) | 1(1,2) | 2.00(1.08,2.76) | 1.79(1.00,2.50) | 29.17(20.00,39.23) |
| Primary schools | 193(48.25) | 13(9,18) | 1(1,3) | 2.10(1.01,3.07) | 2.33(1.25,4.00) | 20.93(13.64,30.21) |
| Junior high schools | 16(4.00) | 11(6,18) | 1(1,4) | 1.96(1.33,3.58) | 2.79(1.92,5.19) | 13.53(8.63,35.67) |
| Senior high schools | 10(2.50) | 15(8,17) | 1(1,8) | 2.39(0.91,4.26) | 4.56(1.63,7.20) | 19.81(5.02,31.77) |
| Others | 16(4.00) | 18.5(15,24) | - | 2.24(2.05,3.84) | 2.61(1.08,5.90) | 15.26(7.85,23.11) |
| Genogroups |  |  |  |  |  |  |
| G I | 53(13.25) | 11(7,13) | 1(1,1) | 2.00(0.78,2.94) | 2.08(0.81,3.50) | 23.19(15.36,32.18) |
| G II | 334(83.50) | 12(9,17) | 1(1,2) | 2.00(1.13,3.00) | 2.00(1.10,3.39) | 24.23(15.69,35.71) |
| G I +G II | 13(3.25) | 13(10,20) | 1(1,3) | 3.13(2.93,3.58) | 2.83(2.39,3.19) | 14.07(10.26,29.70) |
| **Foodborne transmission** | 31(7.18) | 39(27,63) | 16(10,24) | 2.16(1.15,4.50) | 7.17(4.34,9.67) | 3.34(1.64,4.54) |
| Urban-rural |  |  |  |  |  |  |
| Urban | 14(45.16) | 42.5(27,68) | 14(4,22) | 2.57(1.15,4.82) | 6.48(4.29,8.36) | 3.79(1.46,6.41) |
| Rural | 17(54.84) | 34(27,59) | 20(14,24) | 2.10(1.22,4.64) | 8.81(4.02,12.97) | 2.40(1.62,4.10) |
| Settings |  |  |  |  |  |  |
| Kindergartens | 0(0.00) | - | - | - | - | - |
| Primary schools | 11(35.48) | 36(24,63) | 14(10,19) | 2.10(1.08,4.50) | 7.17(3.67,11.42) | 3.66(2.45,5.12) |
| Junior high schools | 1(3.23) | 34(34,34) | 10(10,10) | 0.29(0.29,0.29) | 6.81(6.81,6.81) | 6.29(6.29,6.29) |
| Senior high schools | 13(41.94) | 41(27,72) | 22(16.,25) | 2.83(1.23,6.08) | 9.04(6.47,14.43) | 2.06(1.64,3.50) |
| Others | 6(19.35) | 33.50(22,67) |  | 2.29(1.79,5.55) | 4.26 (2.99,6.75 ) | 4.72(0.60,27.75) |
| Genogroups |  |  |  |  |  |  |
| G I | 5(16.13) | 39(25,62) | 10(3,25) | 2.73(1.69,6.06) | 4.00(3.19,15.42) | 5.64(2.49,16.43) |
| G II | 21(67.74) | 36(28,61) | 15(10,21) | 2.00(1.14,4.37) | 7.16(5.22,8.92) | 2.45(1.62,4.24) |
| G I +G II | 5(16.13) | 51(27,80) | 24(24,26) | 4.32(1.73,6.50) | 14.33(6.85,16.66) | 2.40(0.80,4.22) |
| **Waterborne transmission** | 1(100.00) | 118(118,118) | - | 12.00(12.00,12.00) | 16.70(16.70,16.70) | 0.35(0.35,0.35) |
| **Total** | 432(100.00) | 1(1,2) | 2.04(1.13,3.08) | 2.21(1.17,4.07) | 2.21(1.17,4.07) | 21.31(13.95,31.91) |
| Urban-rural |  |  |  |  |  |  |
| Urban | 267(61.81) | 11(8,17) | 1(1,2) | 2.08(1.17,3.05) | 2.08(1.04,3.46) | 23.68(15.63,35.00) |
| Rural | 165(38.19) | 14(10,21.5) | 2(1,3) | 2.00(1.00,3.42) | 2.67(1.48,6.18) | 20.22(11.01,33.67) |
| Settings |  |  |  |  |  |  |
| Kindergartens | 165(38.19) | 10(8,13.5) | 1(1,1.5) | 2.00(1.08,2.76) | 1.79(1.00,2.50) | 29.17(20.00,39.23) |
| Primary schools | 204(47.22) | 14(9,19.75) | 1(1,3) | 2.10(1.05,3.08) | 2.39(1.30,4.35) | 20.00(12.28,29.48) |
| Junior high schools | 17(3.94) | 12(6.5,19.5) | 1(1,5) | 1.96(1.28,3.52) | 2.83(1.95,5.35) | 12.35(7.48,33.51) |
| Senior high schools | 23(5.32) | 26(15,51) | 15(1,23) | 2.83(1.15,4.96) | 6.90(4.00,9.48) | 3.94(1.87,18.18) |
| Others | 23(5.32) | 20(16,28) | 0(0,3) | 2.25(2.04,4.23) | 3.83(1.92,6.21) | 14.07(5.17,22.22) |
| Genogroups |  |  |  |  |  |  |
| G I | 59(13.66) | 11(7,14) | 1(1,1) | 2.08(0.88,3.46) | 2.33(0.92,3.71) | 21.31(13.95,31.91) |
| G II | 355(82.18) | 12(9,18) | 1(1,2) | 2.00(1.13,3.00) | 2.08(1.16,4.10) | 23.33(13.64,35.00) |
| G I +G II | 18(4.17) | 18.5(11,30) | 2(1,23.25) | 3.24(2.83,4.54) | 3.03(2.73,8.73) | 11.175(4.24,28.72) |

### 3.6 Logistic regression analysis of factors influencing outbreak scale

We categorized clusters into “outbreak” and “non-outbreak” groups, using non-outbreak clusters as controls. Univariate analysis identified significant associations (P<0.05) for transmission route, urban-rural distribution, institutional setting, and pathogen genogroup. Although reporting timeliness was not statistically significant, we included it in the multivariate logistic regression model due to its established epidemiological relevance [3].

The multivariate analysis revealed foodborne transmission carried substantially higher risk of progressing to outbreak status compared to person-to-person transmission (OR=42.13, 95% CI:10.15-174.84). Suburban/rural areas showed 1.78-fold higher outbreak risk than urban areas (95% CI:0.99-3.19). Compared to kindergartens, primary schools had 6.60-fold higher risk (95% CI:2.97-14.74), junior high schools 4.85-fold (95% CI:1.11-21.24), and senior high schools 5.86-fold (95% CI:1.30-26.43). GII genogroup (OR=4.13, 95% CI:1.40-12.22) and mixed infections (OR=6.55, 95% CI:1.23-34.92) showed significantly higher outbreak risk than GI. (See Table 4)

**Table 4.** Analysis of influencing factors on the scale of norovirus lusters in Hangzhou, 2018-2023.

|  | Cluster count | Outbreak count(%) | Univariate analysis |  | Multivariate analysis |  |
| --- | --- | --- | --- | --- | --- | --- |
|  |  |  | OR(95%CI) | P | OR(95%CI) | P |
| **Transmission route** |  |  |  |  |  |  |
| Person-to-person transmission | 400 | 60(15.00) | 1.00 |  | 1.00 |  |
| Foodborne transmission | 31 | 28(90.32) | 52.89(15.59~179.48) | <0.001 | 42.13(10.15~174.84) | <0.001 |
| **Urban-rural** |  |  |  |  |  |  |
| Urban | 267 | 42(15.73) | 1.00 |  | 1.00 |  |
| Rural | 165 | 47(28.48) | 2.13(1.33~3.42) | 0.002 | 1.78 (0.990~3.186) | 0.054 |
| **Settings** |  |  |  |  |  |  |
| Kindergartens | 165 | 8(4.85) | 1.00 |  | 1.00 |  |
| Primary schools | 204 | 51(25.00) | 6.54(3.01~14.24) | <0.001 | 6.598 (2.96~14.74) | <0.001 |
| Junior high schools | 17 | 4(23.53) | 6.04(1.60~22.76) | 0.008 | 4.85 (1.11~21.24) | 0.036 |
| Senior high schools | 23 | 14(60.87) | 30.53(10.18~91.54) | <0.001 | 5.86 (1.30~26.43) | 0.021 |
| Others | 23 | 12(52.17) | 21.401(7.24~63.26) | <0.001 | 10.481 (2.90~37.95) | <0.001 |
| **Genogroups** |  |  |  |  |  |  |
| G I | 59 | 8(13.56) | 1.00 |  | 1.00 |  |
| G II | 355 | 73(20.56) | 1.65(0.75~3.63) | 0.213 | 4.13 (1.40~12.22) | 0.010 |
| G I +G II | 18 | 8(44.44) | 5.10(1.55~16.79) | 0.007 | 6.55 (1.23~34.92) | 0.028 |
| **Reporting timeliness** |  |  |  |  |  |  |
| <1d | 187 | 37(19.79) | 1.00 |  | 1.00 |  |
| 1-3d | 125 | 29(23.20) | 1.23(0.707~2.12) | 0.470 | 1.28 (0.65~2.50) | 0.468 |
| ≥3d | 120 | 23(19.17) | 0.961(0.54~1.72) | 0.894 | 0.51 (0.24~1.091) | 0.082 |

"Outbreak COUNT (%)" indicates the proportion of clusters that met outbreak criteria within each category.

## 4. Discussion

Norovirus currently comprises at least 10 genogroups and 49 genotypes [5]. Multiple studies have shown limited cross-reactive antibody and cross-blocking activity among different genotypes within the GII genogroup [6]. Prior to 2016, the dominant strains circulating in China were GII.4 and GII.17 [7–9]. Since 2016, a new variant, GII.P16-GⅡ.2, has emerged globally [3]. The lack of a robust population immune barrier against this novel strain may explain the significant increase in norovirus clusters in Hangzhou and throughout China between 2018 and 2020. From 2021 to 2023, the implementation of non-pharmaceutical interventions (NPIs) for COVID-19 prevention—such as reduced population mobility, limited gathering sizes, frequent school closures, and shutdowns of restaurants and public venues—along with enhanced hygiene practices including social distancing, mask-wearing, and hand hygiene, likely contributed to the marked decline in norovirus clusters in Hangzhou during this period. Similar trends were observed in other Chinese cities such as Beijing [10] and Yangzhou [11], as well as in countries including the United States [12], Germany [13], and Australia [14].

Compared to other major Chinese cities, Hangzhou reported a higher number of norovirus clusters. Beyond heightened surveillance sensitivity, this may be attributed to several regional factors. As suggested by a study focusing on the Pearl River Delta [15], geographical, climatic, and socioeconomic conditions—as well as local lifestyles—may facilitate norovirus transmission. Hangzhou shares many of these characteristics: situated near the Qiantang River estuary, the city experiences twice-daily tidal surges that push seawater upstream over 100–200 km. This saline intrusion into freshwater aquifers may contribute to norovirus contamination of local nearshore seafood. In addition, Hangzhou is a major commercial and tourist hub, ranking among the top five Chinese cities for annual tourist visits, resulting in high population mobility. Climatically, located at the southern edge of the northern subtropical monsoon zone, the city experiences cool, rainy, and humid conditions in winter and spring (October to May), which correspond to the high-incidence season for norovirus, while clusters are rare during the hot summer months—consistent with the inverse relationship between norovirus survival and temperature [16–17]. Furthermore, very few clusters were reported during the approximately 30-day winter vacation (around January), when schools are closed.

In Hangzhou, norovirus clusters occur primarily in schools, particularly kindergartens and primary schools, aligning with the overall pattern observed across China [3–4]. This concentration can be attributed to several factors. First, schools represent densely populated environments where young children, whose immune systems are still developing and who have not yet fully established habits of personal hygiene, frequently engage in group activities such as playing, off-campus daycare [18], and tutoring—behaviors that increase both susceptibility to initial infection and risk of transmission within the school setting. This is further evidenced by four reported norovirus clusters occurring in off-campus training institutions during the same period, indicating that such facilities may act as transmission hubs.

Secondly, infection control measures in some schools remain suboptimal. Instances of improper handling of vomitus and attendance of symptomatic students who subsequently vomit in classrooms have been reported. Given the high infectivity of norovirus, its rapid transmission dynamics, and the abundance of asymptomatic carriers [4], these conditions significantly contribute to person-to-person spread [19–20]. Clusters have also been recorded in other settings such as hotels, companies, villages, and elderly care facilities, underscoring that any environment characterized by collective dining or group living poses a risk for norovirus clusters.

Notably, epidemiological features vary by educational level. While senior high schools experience fewer overall clusters compared to primary schools, those that do occur are more likely to evolve into large-scale clusters and last longer. This is largely because most high schools in Hangzhou are boarding institutions where all meals are provided centrally onsite, creating heightened risk of foodborne transmission (13 of the 31 foodborne clusters occurred in high schools, accounting for 41.93%). Moreover, due to intense academic pressure, students, parents, and even school authorities often resist class suspensions—one of the most effective control measures [20–21]—and may underreport cases to avoid disrupting instruction, leading to delayed containment and prolonged transmission.The distribution of norovirus outbreak settings in Hangzhou differs from that observed in countries such as the United States. According to U.S. CDC data and related literature, healthcare facilities are the most commonly reported settings for norovirus outbreaks in the U.S. [22–23]. This discrepancy may be attributed to differences in disease surveillance systems, school population densities, aging demographics, elder care practices, and disinfection protocols in medical institutions across countries. Another contributing factor is that Chinese families tend to be highly attentive—sometimes overly so—to children’s symptoms, leading to higher reporting rates in schools. In contrast, adults with diarrhea often do not seek medical care. Surveys indicate that only 31.01% of adults in Hangzhou [24] and 28.4% in Chengdu [25] sought medical attention for diarrhea in 2010, as many consider it a mild condition not requiring professional consultation. This may also contribute to the underreporting of norovirus clusters in non-school settings in China.

In Hangzhou, person-to-person transmission was the dominant route of spread, followed less commonly by foodborne transmission, and rarely by waterborne transmission. This represents a significant shift from earlier patterns. For example, 14 of 16 norovirus outbreaks in Zhejiang Province from 2004 to 2014 were waterborne, and only one was foodborne [26]. Similarly, in Guangdong Province, 40.6% of outbreaks between 2008 and 2015 were foodborne [27]. The marked reduction in foodborne and waterborne transmission likely reflects improvements in public sanitation, hygiene awareness, and food safety management. Moving forward, outbreak control strategies should prioritize interrupting person-to-person transmission through health education, promoting hand hygiene, training teachers in safe vomitus handling, and enhancing disinfection of high-contact surfaces (e.g., door handles, stair railings) in classrooms and dormitories during peak months [28].

Foodborne transmission in school settings can lead to extensive outbreaks and poses a greater public health challenge. Contamination in school cafeterias can expose the entire school population, resulting in a large number of cases that are geographically dispersed and difficult to detect comprehensively. Delayed case isolation further increases the risk of secondary person-to-person transmission. Additionally, foodborne outbreaks occur disproportionately in high schools (13 of 31 foodborne outbreaks, 41.93%). Due to intense pressure associated with China’s national college entrance exams (Gaokao), there is strong resistance among schools, parents, and students to control measures such as case isolation and class suspensions, which are perceived as detrimental to academic performance. This reluctance leads to prolonged outbreaks and higher case numbers compared to person-to-person transmission events.

The occurrence and scale of norovirus clusters are also linked to urban-rural distribution. From 2018 to 2023, 61.81% of clusters occurred in urban areas and 38.19% in suburban/rural regions, likely due to higher population density in urban settings. In 2020, Hangzhou’s urban areas had an average of 3,700 permanent and mobile residents per square kilometer and 0.67 schools (from kindergarten to high school) per km², compared to only 700 people and 0.07 schools per km² in suburban/rural areas. The higher density of people, schools, and interpersonal contact in urban areas may contribute to the increased frequency of clusters. However, 27.32% of clusters in suburban/rural areas progressed to large-scale clusters, compared to 15.73% in urban areas (χ² = 10.142, P = 0.001). This disparity may be due to poorer sanitation conditions—especially in high school cafeterias and dormitories—in suburban/rural schools, increasing the risk of foodborne transmission. Moreover, slower implementation of control measures (e.g., cafeteria closures, professional disinfection due to limited resources or awareness) may facilitate ongoing transmission in these settings. Further detailed investigation is needed to clarify these factors.

China places strong emphasis on the prevention and control of infectious diarrhea in schools. Since 2006, a national surveillance and reporting system for infectious disease symptoms has been established in educational settings. Official guidelines including the *Technical Guidelines for Infectious Disease Prevention and Control in Schools (GB 28932-2012)*, the *Technical Guidelines for Norovirus Outbreak Investigation and Prevention (2015)*, and the *Notice on Issuing Disinfection Guidelines for Norovirus Prevention in Key Settings Including Schools (2024)* have been issued.

Schools in China now implement comprehensive and scientific measures including routine health education, health monitoring, environmental sanitation, and disinfection, as well as case isolation and class suspensions during clusters. These measures have played a crucial role in controlling norovirus transmission.

## 5. Conclusions

The prevention and control of norovirus clusters in Hangzhou should prioritize school settings—especially primary schools and kindergartens—with a focus on interrupting person-to-person transmission, while also addressing the persistent risk of foodborne spread. The following measures are recommended to enhance outbreak management:

1. **Strengthen School-Based Surveillance**: Schools should strictly implement morning and midday health checks, improve the system for tracking illness-related absences, and enhance the sensitivity of outbreak monitoring to ensure timely and complete reporting.
2. **Promote Hygiene and Containment Practices**: Regular training on hand hygiene and standard protocols for safe vomitus disposal should be reinforced [15]. Additionally, daily environmental disinfection should be intensified during peak transmission periods [19].
3. **Regulate Off-Campus Educational Settings**: Enhanced supervision of private tutoring centers and off-campus daycare programs is needed, particularly during the winter and spring. Strict disinfection and symptom reporting protocols must be enforced to reduce cross-infection.
4. **Ensure Food Safety in High Schools**: Canteens in high schools should receive intensified food safety inspections, and regular health monitoring of food service staff should be mandatory to prevent foodborne outbreaks [29].

### Limitations

This study has several limitations. First, China’s current symptom-based infectious disease surveillance and reporting system operates primarily in schools, with limited coverage in other settings such as corporations, restaurants, and nursing homes. Under-reporting in these locations may introduce surveillance bias. Second, due to funding constraints, genotyping of norovirus strains was not performed for every cluster, preventing detailed analysis of local genotype dynamics and specific transmission patterns of individual strains.

## Data Availability

The data underlying the results presented in this study cannot be publicly shared due to ethical restrictions imposed by the Ethical Review Committee of Hangzhou CDC?because the data contain sensitive patient information. However, de-identified data are available upon request to all interested researchers.

## Acknowledgments

We acknowledge the dedicated efforts of all staff members in managing the epidemic response, whose contributions were essential to this investigation.

## Author Contributions

Conceptualization: SONG Kai, DUAN Xiaojian.

Data curation: WANG Jiayun,REN Xiaobin

Formal analysis: Wang Jing, HUO Lianglian

Funding acquisition: HUO Lianglian

Investigation: SONG Kai, DUAN Xiaojian, WANG Jiayun,REN Xiaobin

Methodology: SONG Kai, Wang Jing.

Project administration: DUAN Xiaojian, HUO Lianglian.

Supervision: HUO Lianglian.

Validation: WANG Jiayun.

Visualization: DUAN Xiaojian.

Writing - original draft: SONG Kai.

Writing -review & editing: Wang Jing.

